# Use of finger length ratio as a marker for knee osteoarthritis: A case-control study of 2,456 patients

**DOI:** 10.1101/2020.07.22.20159681

**Authors:** Dukhum Magu, Aditya Aggarwal, Prateek Behera, Ankit Khurana

## Abstract

**Background:** In the human hand, the index and ring finger present a considerable variation in their relative lengths and the ratio of their lengths (2D:4D ratio). This ratio is associated with a variety of behavioral and physiological traits possibly linked to variation in sex hormones levels. Previous studies have revealed inconsistent results while assessing the association of 2D:4D ratio as a risk factor for occurrence of knee osteoarthritis (KOA). This study was designed as a prospective observational study to analyze this association using a better methodology.

**Methods:** Patients were enrolled into KOA group (1396 patients) and non-KOA group (1060 patients) based on ACR criteria for OA knee. Knee and hand radiographs of all patients enrolled were assessed. The 2D:4D length ratio was calculated for phalanges, metacarpal bones and for the combined (metacarpal & phalanx) finger lengths on radiographs and visual finger lengths. The finger patterns were classified and assessment was done between these ratios and finger pattern types for occurrence and severity of KOA.

**Results:** A lower 2D:4D ratio in an individual was associated with an increased chance of KOA and a dose response relationship was found between radiographic grading of KOA and 2D:4D ratio.

**Conclusion:** Based on the findings of this study one can predict the risk of developing KOA using this simple technique which can be used as a screening tool whereby preventive intervention can be started if someone presents early with a low 2D:4D finger length ratio.

**KEY MESSAGES:**

- Evaluation of finger patterns reveals an association with osteoartritis knee (KOA)
- Low 2D:4D ratio and Type 3 finger pattern is associated with increased KOA risk
- Low 2D:4D ratio has as positive correlation to radiographic severity of KOA
- Preventive interventons can be initiated for those with at risk finger pattern

## Introduction

Osteoarthritis (OA) is among the most prevalent diseases in the world.(1) It is one of the leading causes of disability among elderly. Numerous risk factors are suspected to play a role in the causation of primary OA. They can be broadly classified as non-modifiable and modifiable. While the non-modifiable risk factors for OA are age, sex, genetic influences and race; modifiable risk factors include occupation, nutrition, muscle weakness, knee instability, malalignment and abnormal joint loading (including obesity).(2,3) It has been suggested that estrogen depletion plays a role in the onset and progression of OA. Men are known to have a higher prevalence of OA than women before the age of 50 (4), but after this age the prevalence is higher in women.(5,6) The prevalence increases with age in both men and women, but in women, it increases dramatically after the age of 50 (7), which coincides with menopause.

In the human hand, the 2^nd^ (index finger) and the 4^th^ (ring finger) digits present a pattern of approximate symmetry around the central axis of the 3^rd^ (Middle finger) digit. However, there is considerable variation in the ratio of the lengths of the 2^nd^ to the 4^th^ digit (2D:4D). Many individuals have a longer 2^nd^ digit (2D:4D>1) while many have a longer 4^th^ digit (2D:4D<1). The former is usually more common in females and the latter in males. (8) The index to ring finger length ratio (2D:4D) is believed to be associated with a wide variety of behavioral and physiological traits. (8– 12) Finger length patterns have been studied in relation to several diseases and physiologic traits and have been linked to the levels of sex hormones. Associations have been described with coronary artery disease, autism, infertility, and age at menarche. Based on the 2D:4D ratio, the hand has been classified into three types - index finger longer than ring finger (type 1), equal to the ring finger (type 2), or shorter than the ring finger (type 3). (8–12) Men are over 2.5 times more likely than women to have a type 3 pattern. Type 3 finger pattern was also associated with a female estrogen deficiency and could be considered a surrogate marker of earlier onset of menopause. (13)

The association of 2D:4D ratio has been assessed as an independent risk factor to occurrence and severity of knee osteoarthritis (KOA) in few previous studies.(13–16) However, the results are inconsistent among them. Their reported inconsistencies are probably related to ethnicity of their study population or are from a difference in the case definition of osteoarthritis considered by them. Our aim was thus to assess whether predicting the possibility of developing a future knee osteoarthritis could be based on this association using a better methodology. This has the advantage of early initiation of preventive strategies thereby reducing the burden of this morbid disease.

## Methods

This prospective observational study was performed at a tertiary care teaching hospital in Northern India. Institutional ethical committee approval was obtained prior to initiation of the study. All the patients reporting to the out-patient clinic of the Department of Orthopedics over a 2 years period (2014-2016) were considered and those who consented were included. Two groups of patients were enrolled – those with KOA (KOA group) and those without (non-KOA group). Inclusion criteria for the KOA group were age more than 25 years and a diagnosis of primary KOA based on American college of Rheumatology criteria (ACR criteria) (17) irrespective of the grade of OA (Kellgren - Lawrence grade). Patients were excluded if they had KOA secondary to trauma, inflammatory diseases (rheumatoid arthritis, ankylosing spondylitis etc.) or as a sequelae of infective diseases like septic arthritis. Patients with crystal arthropathy and with any injury or deformity of the hand not from primary OA were also excluded. The non-KOA patients were also >25 years of age and had presented to the clinic with complaints other than OA.

Demographic characteristics with a potential to cause or contribute to KOA were recorded in a pre-defined format and included weight, height, waist circumference (WC), blood pressure and history of other risk factors like smoking, alcohol intake, diabetes mellitus and hypertension. Postero-anterior (PA) radiographs of the hands were obtained. Finger length measurements were obtained for participants of both the groups by two methods – visual and radiographic. For visual finger length the soft tissue outlines of the index and ring finger were measured from the base of a digit to its tip using a Vernier caliper. The Vernier caliper is an extremely precise measuring instrument; the reading error is 1/20 mm = 0.05 mm. The radiographic method involved measuring the lengths of 2^nd^ (2D) & 4^th^ metacarpal (4D) and 2^nd^ (2D) & 4^th^ phalanx (4D) with on 100% magnification X-rays of the hand by an independent observer and the 2D:4D length ratio was calculated. (Figure 1) The radiographic phalanx length is the length from the midpoint of the base of the proximal phalanx to the midpoint of the tip of the distal phalanx, and the radiographic metacarpal length (cm) is measured from the midpoint of the base of the metacarpal to the midpoint of the tip of the metacarpal bone. The 2D:4D length ratio was calculated separately for phalanges and metacarpal bones, as well as for the combined (metacarpal & phalanx) finger lengths. This has been described in Table 1. The values for left and right hand were averaged out.

**Table 1:** Parameters of 2^nd^ and 4^th^ digits studied and compared to osteoarthritis of the knee.

| Measurement | Definition |
| --- | --- |
| Radiographic Phalanx length (in cm) | Length from midpoint of the base of the proximal phalanx to the midpoint of the tip of the distal phalanx on 100% PA view |
| Radiographic Metacarpal length (in cm) | Length from midpoint of the base of the metacarpal to the midpoint of the tip of the metacarpal bone |
| Radiographic combined length (in cm) | Radiographic Phalanx length + Radiographic metacarpal length |
| Visual finger length (in cm) | Clinically measured length from base of a digit to apex using a vernier calliper |
| 2D:4D phalangeal ratio | Radiographic Phalanx length of 2<sup>nd</sup> digit divided by Radiographic Phalanx length of 4<sup>th</sup> digit |
| 2D:4D metacarpal ratio | Radiographic metacarpal length of 2<sup>nd</sup> digit divided by Radiographic metacarpal length of 4<sup>th</sup> digit |
| 2D:4D combined ratio | Radiographic combined length of 2<sup>nd</sup> digit divided by Radiographic combined length of 4<sup>th</sup> digit |
| 2D:4D visual assessed ratio | Visual finger length of 2<sup>nd</sup> digit divided by visual finger length of 4<sup>th</sup> digit |

**Figure 1:**
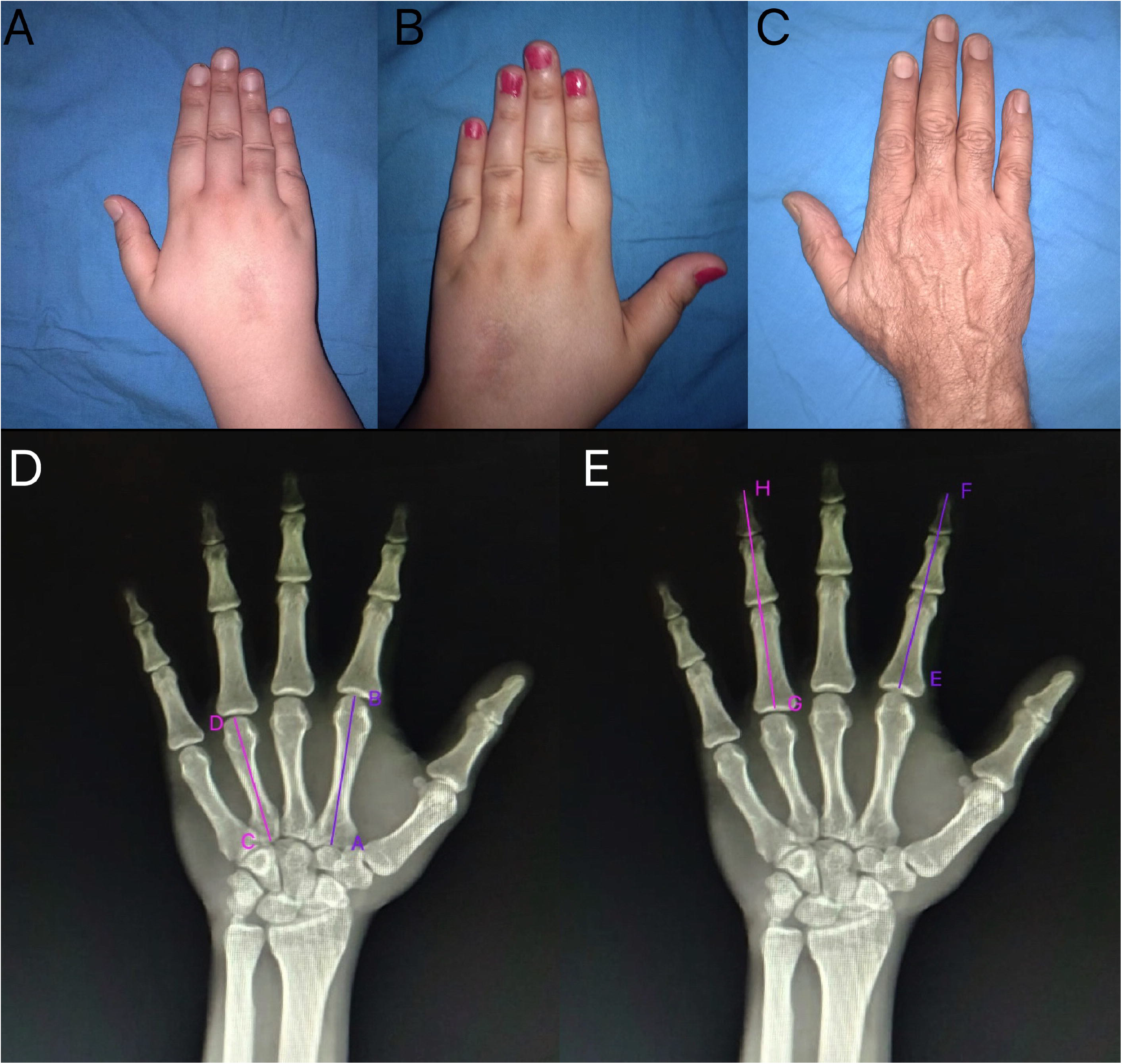
(A, B and C) Representing Type 1, 2 and 3 finger pattern types as assessed visually. (D) PA radiographs of hand showing measurement of radiographic 2D:4D metacarpal ratio (*ab/cd*); (E) PA radiographs of hand showing measurement of radiographic 2D:4D phalanx ratio (*ef/gh*)

Average finger length was classified into 3 types based on relative length of the index finger with respect to ring finger and index finger was either longer (type 1), equal to (type 2), or shorter than the ring finger (type 3). (Figure 1 A,B and C) All patients (both KOA and non-KOA groups) underwent standing knee radiographs to confirm the diagnosis of KOA and to assess for KL grading. In cases with bilateral OA knee the side with higher grading was used for evaluation. In case a patient in the non-KOA group had any evidence of OA on radiograph, he/she was excluded from analysis.

The measurement of lengths of the digits on the radiographs and the data analysis were done by researchers other than the one involved in measurement of lengths of the digits on the patients. Data were coded and recorded in MS Excel spreadsheet program. R Statistical Software (R Core Team, Vienna, 2019) was used for data analysis. Group comparisons for continuously distributed data were made using independent sample ‘t’ test. Chi-square test was used for group comparisons for categorical data. In case the expected frequency in the contingency tables was found to be <5 for >25% of the cells, Fisher’s Exact test was used instead. Linear correlation between ordinal/continuous data was explored using Spearman’s rank correlation. Univariable and multivariable binary logistic regression was performed with group as the dependent variable to explore the association of various 2D:4D ratios with KOA, while controlling for age, gender, and BMI. Statistical significance was kept at p < 0.05.

## Results

1396 patients satisfying the inclusion and exclusion criteria were included in KOA group. Additionally, 1060 healthy individuals were enrolled in the non-KOA Group. All 2456 patients had their radiographic metacarpal and phalanges ratios measured. The combined length of the metacarpal and the phalanges were measured by adding the individual lengths. As no difference was observed between the measurements in the right and the left hands, values for both right and left hands were averaged to obtain the final values used for analysis.

Mean age of individuals in the KOA group was 63.7 years while that of non-KOA group was 49.5 years. Majority of the patients were females (1017; 72.9%), while the rest (379; 27.1%) were males. Further analysis of sex distribution among different age categories showed that the largest proportion of females (34.8%) were in the age group of 56-65 years and the largest proportion of males (31.2%) were in the age group of 46-55 years. 71% of the KOA patients had bilateral disease. Most patients (70.5%) had BMI ≥ 25 kg/m^2^ and hence belonged to overweight-obese category. The maximum BMI noted was 50 kg/m^2^ in the KOA group with a mean BMI of 29.9 kg/m^2^. In the non-KOA group, the maximum BMI noted was 45 kg/m^2^ with a mean BMI of 25.9 kg/m^2^. The distribution of smokers and alcoholics were comparable among both the groups.

The distribution of the different finger patterns and the mean values of measured 2D:4D length ratios are summarized in Table 2. Comparison of the different types was done based on the radiographic metacarpal length, radiographic phalanx length, radiographic combined length and visual finger length. Radiographically, type 1 pattern was the commonest pattern observed in both KOA and non-KOA groups, but there were more OA patients who had type 3 pattern (33 vs 0) and this difference was statistically significant (p<0.001). A similar pattern was also observed in radiographic phalanx lengths (13 vs 0; p<0.001). Analysis of the combined radiographic lengths of both phalanx and metacarpal revealed that like the other ratios, here too type 1 was the commonest type in both KOA and non-KOA groups, and type 3 was significantly more common in KOA group as compared to the non-KOA group (p=0.042). The visually assessed finger length pattern showed that type 3 pattern was the commonest in both groups, and that a significantly larger proportion of patients with KOA had this pattern, as compared to the non-KOA (p<0.001). Thus, when compared with other finger pattern types (types 1 and 2), the type 3 finger pattern was seen more frequently in KOA patients in terms of visual and all radiographic parameters studied.

**Table 2:**
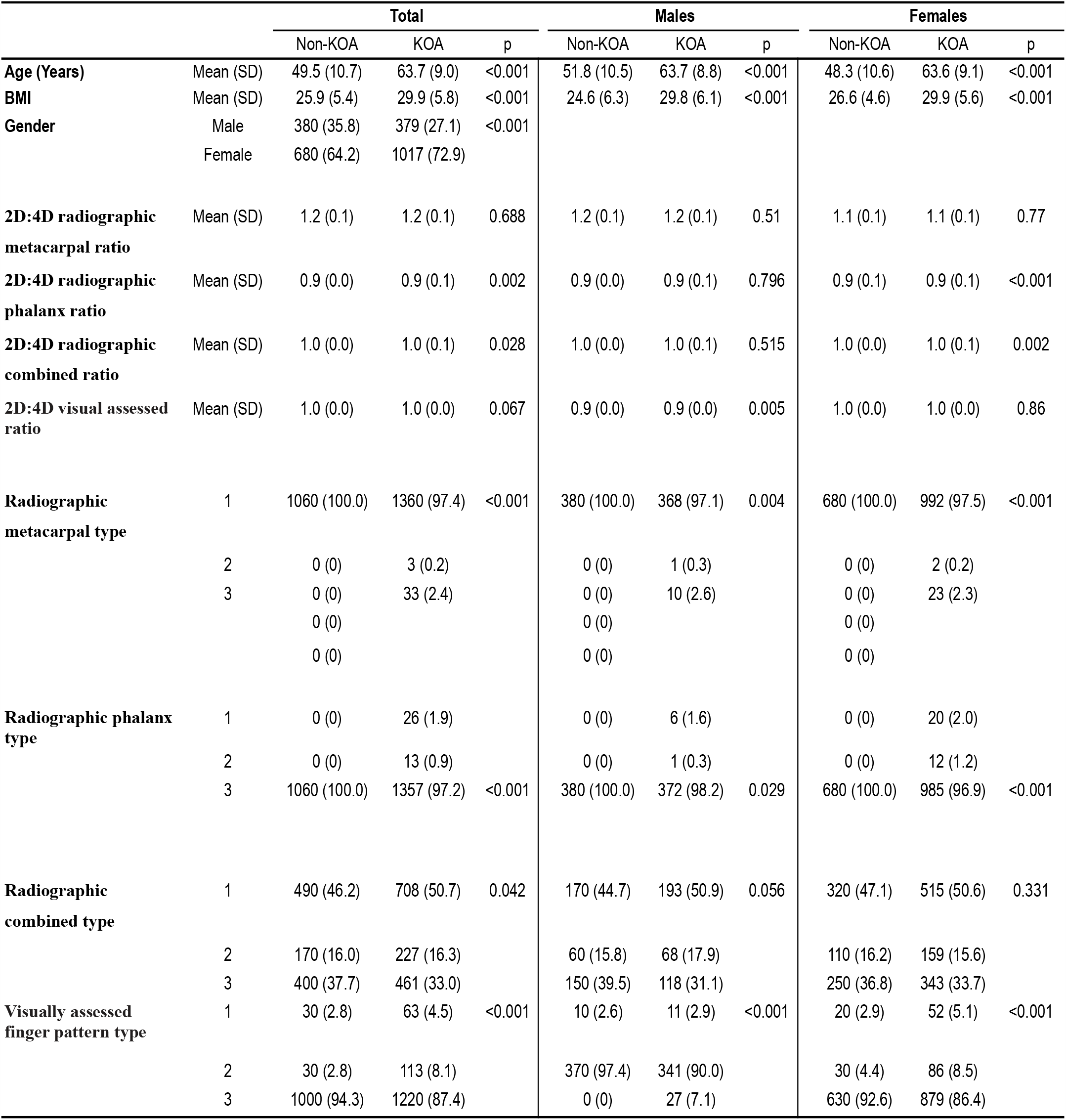
Characteristics of study population including digital measurements and their distribution between OA and non-OA groups.

In univariable binary logistic regression with group as dependent variable, significant odds ratios were observed for age (OR=1.15, p<0.001), BMI (OR=1.15, p<0.001) and gender: female (OR=1.50, p<0.001), thus suggesting that increasing age & BMI, and female gender predisposed to KOA. We then performed multivariable binary logistic regression with group as the dependent variable, each of the 4 ratios one-by-one as the predictor variables, and age, BMI and gender as covariates in each of the 4 models. It was seen that a lower 2D:4D radiographic metacarpal ratio lead to higher odds of KOA (OR=0.03, p<0.001), with a decrease of 0.1 in this ratio increasing the odds of KOA by 9.7%. When this analysis was performed separately in males and females, a similar association was observed. Additionally, in males, it was observed that a decrease in 2D:4D radiographic combined ratio also increased the odds of KOA. (Table 3)

**Table 3:** Odds Ratios of 2D:4D Ratios Controlling for Baseline Characteristics

| 2D:4D Ratios | Odds Ratios of 2D:4D Ratios Controlling for Baseline Characteristics |  |  |
| --- | --- | --- | --- |
|  | Full Sample | Males | Females |
| 2D:4D radiographic metacarpal ratio | 0.03 (0.01-0.14, p<0.001) | 0.05 (0.01-0.32, p=0.002) | 0.02 (0.00-0.14, p<0.001) |
| 2D:4D radiographic phalanx ratio | 1.08 (0.12-10.54, p=0.947) | 6.12 (0.10-566.82, p=0.405) | 0.34 (0.03-5.08, p=0.423) |
| 2D:4D radiographic combined ratio | 0.14 (0.02-0.99, p=0.056) | 0.01 (0.00-0.43, p=0.015) | 0.31 (0.03-3.10, p=0.329) |
| 2D:4D visual assessed ratio | 13.71 (0.38-516.61, p=0.155) | 937.69 (1.52-624767.41, p=0.038) | 2.37 (0.03-193.12, p=0.698) |

Most of the patients in the KOA group had KL Grade 1 and 2 OA (434/1396 and 705/1396 respectively). Correlation analysis between KL grade and 2D:4D ratios and finger types showed that the phalangeal ratio and combined radiographic digital length ratio & type showed significant negative correlation with KL grade. The correlation was however weak (spearman rho<0.3). This has been summarized in table 4.

**Table 4:** Correlation of digital measurements (2D:4D ratio) and type of digit with KL grade

| Correlation of KL grade with: | Spearman's Rho | P value | Inference |
| --- | --- | --- | --- |
| 2D:4D radiographic metacarpal ratio | -.034 | .203 | Insignificant (p>0.05) |
| Radiographic metacarpal type | -.007 | .781 | Insignificant (p>0.05) |
| 2D:4D radiographic phalanx ratio | -.064 | .017 | Significant, Poor |
| Radiographic phalanx type | -.002 | .947 | Insignificant (p>0.05) |
| 2D:4D radiographic combined length ratio | -.071 | .008 | Significant, Poor |
| Radiographic combined length type | .073 | .006 | Significant, Poor |
| 2D:4D visual assessed ratio | .020 | .447 | Insignificant (p>0.05) |
| Visually assessed finger pattern type | -.051 | 0.056 | Insignificant (p>0.05) |

## Discussion

Osteoarthritis (OA) is presently considered as the end result of a molecular cascade which ensues after certain triggers occur and ultimately results in irreversible damage to the articular cartilage. (18) In this study, a unique association that has been sparingly studied earlier was examined. Present study found that a lower 2D:4D ratio in an individual was associated with an increased chance of osteoarthritis of the knee. Not only did we find an association with prevalence of osteoarthritis we also found a dose response relationship between radiographic grading of osteoarthritis of the knee and 2D:4D ratio. Our findings were consistent with Zhang et al, Ferraro et al and Hussain et al. (13,14,19) The difference in OA risk in relation to 2D:4D may be explained, at least in part, by the different susceptibility of knee in response to injury. Hormonal changes are believed to be associated with altered 2D:4D ratio. Hormonal influences can affect growth of bones, cartilage and soft tissues. Population with lower 2D:4D ratio have been proposed to have higher testosterone levels.(20–22) This also in part explains the increased incidence of KOA in females after the age of 50 (post menopause) and in males under the age of 50. It has also been reported that a lower 2D:4D ratio which is associated with higher androgen levels may lower the pain thresholds thereby leading to more pain for the same grade of OA changes. (23)

Previous studies which have examined the association between finger length type and OA risk had inconsistent findings. The inconsistencies could be due to a difference in the working definition of OA used in them. Zhang et al and Ferraro et al used a clinical definition of OA and reported an association between finger pattern and OA risk.(13,14) Haugen et al did a radiographic analysis and found no similar association.(16) De Kruijf et al hypothesized that the associations reported might have been driven by pain rather than joint damage which means that that increased pain susceptibility rather than a clear association with joint damage was linked to the associations reported. We however took a combined definition of OA. (23) Patients who satisfied the clinical ACR criteria for OA knee and had radiologic OA were included in the KOA group. (17) Patients in non-KOA group were neither symptomatic nor had radiologic OA thereby making our study better.

Some authors have used joint replacement as an indicator for osteoarthritis.(13,19) This corollary probably cannot be used in Asian/Indian patients as most of them present late and have a general tendency to delay surgery if possible. The ACR criteria used to diagnose osteoarthritis and Kellgren Lawrence grading for radiographically grading OA gave us a more objective diagnostic measure than joint replacement rates. We believe that undergoing a TKR is not just a medical decision but also a financial and social decision especially in counties where patient pays out of pocket for treatment. Muraki et al showed that the odds ratio for knee pain of KL ≥ 3 OA was much higher than that of KL≥ 2 OA in both genders, suggesting that joint space narrowing was more strongly associated with the pain than osteophytosis.(24) This supports our use of KL grade as an indicator of severity of knee osteoarthritis.

Ferraro and Sigurjonsdottir et al (14,15) did not consider if their participants had hand OA, which might be an important confounding factor. We have excluded cases with deformity or/and OA of the hand as these findings can potentially alter the 2D:4D ratio. Considering the same hypothesis, patients with altered 2D:4D ratio also have a higher tendency to develop OA of the hand as a part of generalized OA. Moreover, OA of the fingers can lead to shortening because of narrowing of the joint space from cartilage breakdown and can therefore confound the finger length pattern and lead to misclassification.

To further substantiate our findings, radiographic measurements of phalanx, metacarpal, combined lengths and visual digit length were obtained and compared individually to occurrence and radiographic grade of KOA. Our findings suggest that a shorter 2^nd^ digit or a decreased 2D:4D ratio which corresponds to type 3 digits is associated with KOA. We also found that metacarpal of the 2^nd^ digit is longer that 4^th^ in majority of the KOA and non-KOA group patients but the ratio tends to decrease in KOA group than the non-KOA group. The phalangeal length and visual digital lengths show 2^nd^ digit to be shorter than the 4^th^ in both groups and the length of 2^nd^ digit is further decreased in KOA group and with increasing severity (KL grade) of KOA although the correlation is weak. To our knowledge, this is the first study to assess all possible digital (visual and radiographic) measurements and correlate these findings to radiographic severity of KOA.

Another strength of the present study was the reasonable size of population-based cohort. Visual length measurements were made with a high degree of reliability using of a Vernier caliper.(25) Use of strict inclusion criteria and exclusion criteria is also a strength of this study. However, there are few limitations. Our findings may represent a selection bias as the patient population had a high BMI, a mean age above 40 years with most being community ambulators belonging to a single ethnic group (North Indians). Measurements were done by one person and non-analysis of intra and inter-observer variations are major limitations. The study could have been made better by making it multi-centric with institutes catering to population in different regions of the country.

In conclusion, a lower 2D:4D ratio and type 3 finger length pattern was found to be associated in middle age and elderly people with an increased risk of KOA. Additionally, it was found that the lower the 2D:4D phalangeal ratio, the higher was the KL grade although the statistical correlation with KL grade was weak. The etiopathogenesis of these findings warrants further investigations. Based on the present study, it can be stated that one can predict the risk of an individual developing KOA by measurement of 2D:4D ratio. If a person in the 3^rd^ to 4^th^ decade of life comes in contact with a clinician for any reason and while taking routine anthropometric measurements (height and weight), the ratio of finger lengths can be calculated to stratify if he/she has increased chance of developing KOA. Following which, the patient can be referred preventive interventions for KOA. This could potentially to reduce the burden of this morbid condition that widely affects middle age and elderly population worldwide.

## Data Availability

All relevant data referred to in the manuscript is available with the corresponding author

## Disclosures

### ETHICS

Institute ethical approval reference number NK/1390/MS/2767 Date of aprooval 2nd April 2014.

